# Prevalence and Clinical Significance of Adult-Onset Cancer Predisposition Variants in Pediatric Oncology

**DOI:** 10.64898/2026.06.07.26354365

**Authors:** Jamie L. Maciaszek, Victor Pastor Loyola, Taylor Cain, Maria Cardenas, Patrick R. Blackburn, Mark R. Wilkinson, Selene C. Koo, Chih-Hsuan Wu, Cai Li, Lu Wang, Kim E. Nichols, Jeffery M. Klco, Mohammad K. Eldomery

## Abstract

**Purpose:** Pathogenic or likely pathogenic (P/LP) variants are increasingly identified in genes more commonly associated with adult-onset cancer predisposition, but their prevalence and relevance to a child’s presenting cancer remain unclear.

**Methods:** We retrospectively analyzed 1,280 consecutive pediatric patients with cancer who underwent clinical germline sequencing, using a virtual panel, from 2021 to 2024. Genes with P/LP variants were categorized as aoCPG or pediatric-onset cancer predisposition genes (poCPG) according to cancer risk before age 18 years and pediatric surveillance recommendations. Variant relevance was adjudicated using tumor diagnosis/histopathology, immunohistochemistry, and tumor molecular features and classified as primary, secondary, or indeterminate.

**Results:** Among 1,280 patients, 197 (15.4%) harbored 211 P/LP variants across 54 genes. Sixty-six variants (31.3%) occurred in aoCPG, 87 (41.2%) in poCPG, and 58 (27.5%) were heterozygous variants in autosomal recessive genes. Among adult-onset variants, 7 (10.6%) were primary, 54 (81.8%) secondary, and 5 (7.6%) indeterminate. Among pediatric-onset variants, 77 (88.5%) were primary and 10 (11.5%) secondary. Six patients (3 adult-onset variants; 3 pediatric-onset variants) received targeted therapy informed by germline/somatic sequencing results.

**Conclusion:** In pediatric oncology, most variants in aoCPG are secondary rather than tumor-related findings. Tumor-informed interpretation, beyond variant classification, may improve reporting, counseling, and therapeutic decision-making

## Introduction

Hereditary cancer predisposition syndromes (CPS) in pediatric patients are commonly driven by highly penetrant genes—here referred to as pediatric-onset cancer predisposition genes (poCPG)—that confer a 10- to 100-fold or greater increase in relative cancer risk during childhood.^1^ However, the widespread adoption of next-generation sequencing and broad panel testing has led to increased detection of variants in so-called adult-onset cancer predisposition genes (aoCPG) in children with cancer.^1–3^ The determination of whether pathogenic/likely pathogenic (P/LP) variants in these genes contribute directly to tumor development may influence clinical trial eligibility and access to targeted therapies, while also informing genetic counseling of the patient and other family members.^4–7^

Several seminal studies have underscored the importance of integrating somatic and germline data when evaluating CPS in pediatric oncology to accurately define the role of a variant in tumorigenesis.^8–12^ In parallel, the American College of Medical Genetics and Genomics Secondary Findings Working Group (ACMG-SFWG) primarily recommends the use of clinical test indication and diagnosis to determine the role and relevance of germline variants identified through germline genome sequencing (WGS) or exome sequencing (WES). Specifically, the ACMG-SFWG framework designates P/LP variants that are relevant to the test indication or diagnosis as primary findings (PF).^13,14^ The ACMG-SFWG recommends the use of secondary finding (SF) terminology when reporting P/LP variants identified through germline WGS or WES that are unrelated to the test indication but occur in a defined subset of highly penetrant genes.^15^

In this study, we analyzed clinical germline and tumor sequencing data from 1,280 consecutive pediatric patients with cancer at our institution (2021–2024) to (1) characterize the clinical and molecular features of aoCPG compared with poCPG, and (2) assess the feasibility of integrating tumor diagnosis/histopathology, and somatic profiling to determine the role of the variant and inform the application of PF and SF terminology.

## Methods

### Study Design and Cohort Assembly

This retrospective cohort study included consecutive pediatric patients with cancer who underwent clinical germline sequencing at St Jude Children’s Research Hospital from 2021 to 2024. Patients were identified through routine clinical testing during treatment or consultation. Because all eligible patients were included, no sample size calculation was performed. Outcomes were based on available clinical, pathologic, and molecular data at the time of review. Germline and tumor sequencing were performed under institutional clinical consent, and the institutional review board approved the study with a waiver of informed consent.

### Sequencing and Variant Curation

Dual germline WGS and WES were performed on peripheral blood or skin samples in a CLIA-certified laboratory, as previously described.^16^ Of the 1,280 patients who underwent germline sequencing, 989 also underwent tumor sequencing using comprehensive DNA/RNA approaches: triple-platform genome, exome, and transcriptome sequencing (n=652); exome plus RNA sequencing (n=293); or RNA sequencing alone (n=44). Tumor panel sequencing results were reviewed when available. Sequence alignment to GRCh37, variant calling, and bioinformatic analyses were performed as previously described.^16^ A 115-gene or 102-gene virtual panel was evaluated depending on testing date (Supplementary Table 1). Germline variants were classified per ACMG and ClinGen recommendations.^17,18^

### Classification of Adult-Onset and Pediatric-Onset Cancer Predisposition Genes

In the absence of a consensus aoCPG definition, we performed a literature review and categorized genes harboring P/LP variants as aoCPG or poCPG based on reported ages of disease manifestation, cumulative cancer risk, and availability of pediatric surveillance recommendations.^1,4,19^ Genes were categorized as aoCPG if they showed high or moderate penetrance in adulthood, conferred low-to-moderate cancer risk (<5%) before age 18, and lacked pediatric surveillance recommendations (Figure 1; Supplementary Table 1). For instance, the aoCPG included heterozygous P/LP variants in *PALB2* and *BRCA2,* which are associated with medulloblastoma; *BRCA2* associated with rhabdomyosarcoma or childhood non–Hodgkin lymphoma; heterozygous DNA mismatch repair (MMR) variants associated with Lynch syndrome; and *BRCA1*, *BARD1*, and *RAD51C* that are associated with neuroblastoma.^1,3,19–21^ In *APC*, only the founder variant NM_000038.6:c.3920T>A p.(Ile1307Lys) was classified as aoCPG.^22^ Overall, the aoCPG gene set comprised heterozygous P/LP variants in autosomal dominant genes, including *APC* p.(Ile1307Lys), *ATM*, *AXIN2*, *BARD1*, *BRCA1*, *BRCA2*, *BRIP1*, *CDKN1B*, *CDKN2A*, *CHEK2*, *DDX41*, *FLCN*, *MITF*, *MLH1*, *MSH2*, *MSH6*, *PALB2*, *PMS2*, *POT1*, *RAD51C*, and *RAD51D*.

**Figure 1.**
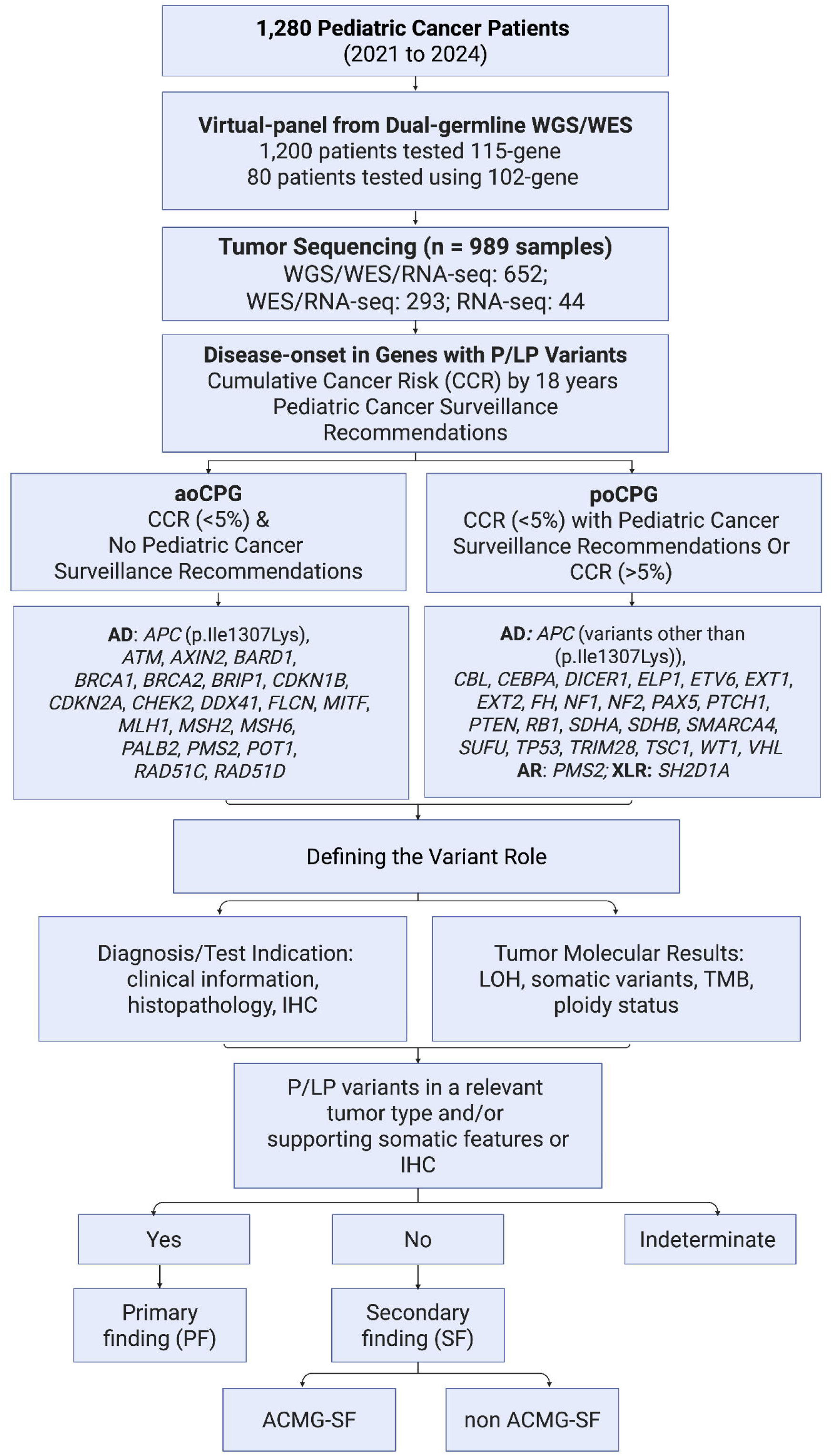
Workflow for defining the role of pathogenic/likely pathogenic (P/LP) germline variants in a pediatric cancer cohort. Flow diagram summarizing the evaluation of 1280 pediatric patients from 2021 to 2024, including 945 tumors with DNA-based tumor sequencing (WES and/or WGS) and an additional 44 tumors with RNA sequencing only. A total of 211 P/LP germline variants were identified in 197 patients using either a 115-gene or 102-gene virtual panel based on the time of testing. Genes were categorized based on cumulative cancer risk by 18 years and the presence of pediatric surveillance recommendations to distinguish adult-onset cancer predisposition genes (aoCPGs) from pediatric-onset cancer predisposition genes (poCPGs). Variant relevance to the presenting tumor was assessed by incorporating the clinical features, tumor diagnosis, histopathology, immunohistochemistry, and tumor molecular findings. Final designation included primary findings (PF), secondary findings (ACMG-SF and non-ACMG-SF), and indeterminate findings. Heterozygote P/LP variants in autosomal recessive genes are not shown. Abbreviations: AD: autosomal dominant; AR: autosomal recessive; XLR: X-linked recessive.

The poCPG category comprised genes with well-established cancer manifestations during childhood (ages 0–18 years), including those with low-to-moderate cancer risk (<5%) but pediatric surveillance recommendations (e.g., *FH*, *NF2*, *PTEN*), high risk (5–50%, e.g., *WT1*), or very high risk (>50%, e.g., *RB1*) (Figure 1).^19,23–27^ *ELP1*, which is associated with Sonic Hedgehog medulloblastoma – a tumor presenting in the first decade of life – carries low-to-moderate (<5%) cancer risk and was classified as poCPG.^28^ This group included autosomal dominant *APC* (variants other than p.(Ile1307Lys)), *CBL*, *CEBPA*, *DICER1*, *ELP1*, *ETV6*, *EXT1*, *EXT2*, *FH*, *NF1*, *NF2*, *PAX5*, *PTCH1*, *PTEN, RB1*, *SDHA*, *SDHB*, *SMARCA4*, *SUFU*, *TP53*, *TRIM28*, *TSC1*, *WT1*, and *VHL*; X-linked recessive *SH2D1A*; biallelic P/LP variants in *BRCA2*, *MLH1*, *MSH2*, *MSH6*, *PALB2*, and *PMS2*. The complete poCPG list appears in Figure 1. Patients with heterozygous variants in autosomal recessive genes were evaluated separately from aoCPG and poCPG.

### Clinical, Histopathologic, and Molecular Review

Clinical data, imaging, tumor histopathology, immunohistochemistry, and tumor genomic data were reviewed for patients with P/LP germline variants in aoCPG or poCPG. Age was defined at diagnosis; sex was abstracted from the medical record; race was self-reported and recorded in the electronic health record. Family history was self-reported during cancer predisposition clinic visits; a positive family history was defined as at least one first- or second-degree relative with cancer diagnosed before age 50, excluding cervical cancer and nonmelanoma skin cancer.^11^

A second somatic event was defined as loss-of-heterozygosity (LOH), a second somatic variant, or absent protein expression by immunohistochemistry.^29^ LOH, used as a surrogate for somatic copy-number loss or copy-neutral LOH, was defined as a tumor variant allele fraction increase of ≥20%, corroborated by tumor WGS-derived copy-number alterations when available.^30^ Tumor mutational burden was categorized as hypermutated (5-100 mut/Mb) or ultrahypermutated (>100 mut/Mb).^5,31^

### Determining the Variant Role

Variants were classified as PF when detected in a relevant tumor type with an established disease–gene association and/or supported by concordant somatic features as defined above. P/LP variants in CPGs unrelated to the tumor type and lacking supporting molecular evidence were classified as ACMG-SF if the gene appeared on the ACMG-SFWG v3.3 gene list,^32^ or as non-ACMG SF if not.^33^ Variants for which the contribution to tumorigenesis could not be determined due to insufficient data were classified as indeterminate.

### Statistical Methods

A variant-level analysis compared the distribution of PF, ACMG-SF, non-ACMG-SF, and indeterminate findings between aoCPG and poCPG using Fisher’s exact test. A secondary patient-level analysis, restricted to patients with autosomal dominant aoCPG or poCPG variants who underwent triple-platform or exome plus RNA sequencing, compared the frequency of second somatic events using Pearson’s Chi-squared test with Yates continuity correction. Because some patients harbored more than one P/LP variant and second-hit evidence contributed to variant adjudication, inferential analyses were interpreted as exploratory. Missing data were not imputed; analyses were performed using available data, with denominators reported where data were incomplete. A 2-sided P<.05 was considered nominally significant. Analyses were performed using R version 4.4.2 (R Foundation for Statistical Computing).

## Results

### Cohort Characteristics and Overall Diagnostic Yield

The cohort included 680 male patients (53.1%) and 600 female patients (46.9%), with a median age at diagnosis of 7 years. Among 1280 patients, 197 (15.4%) harbored germline P/LP variants, including 65 (5.1%) with at least 1 aoCPG variant, 86 (6.7%) with at least 1 poCPG variant, and 57 (4.5%) with at least 1 heterozygous variant in an autosomal recessive gene. Ten patients had variants in more than 1 gene category (Table 1). In total, 211 P/LP variants were identified across 54 genes, including 66 (31.3%) in aoCPG, 87 (41.2%) in poCPG, and 58 (27.5%) heterozygous variants in autosomal recessive genes. Tumor sequencing data were available for 151 of 197 variant-positive patients (77%), including 98 analyzed by triple-platform sequencing, 49 by exome plus RNA sequencing, 7 by targeted panels and 4 by RNA-seq only.

**Table 1.**
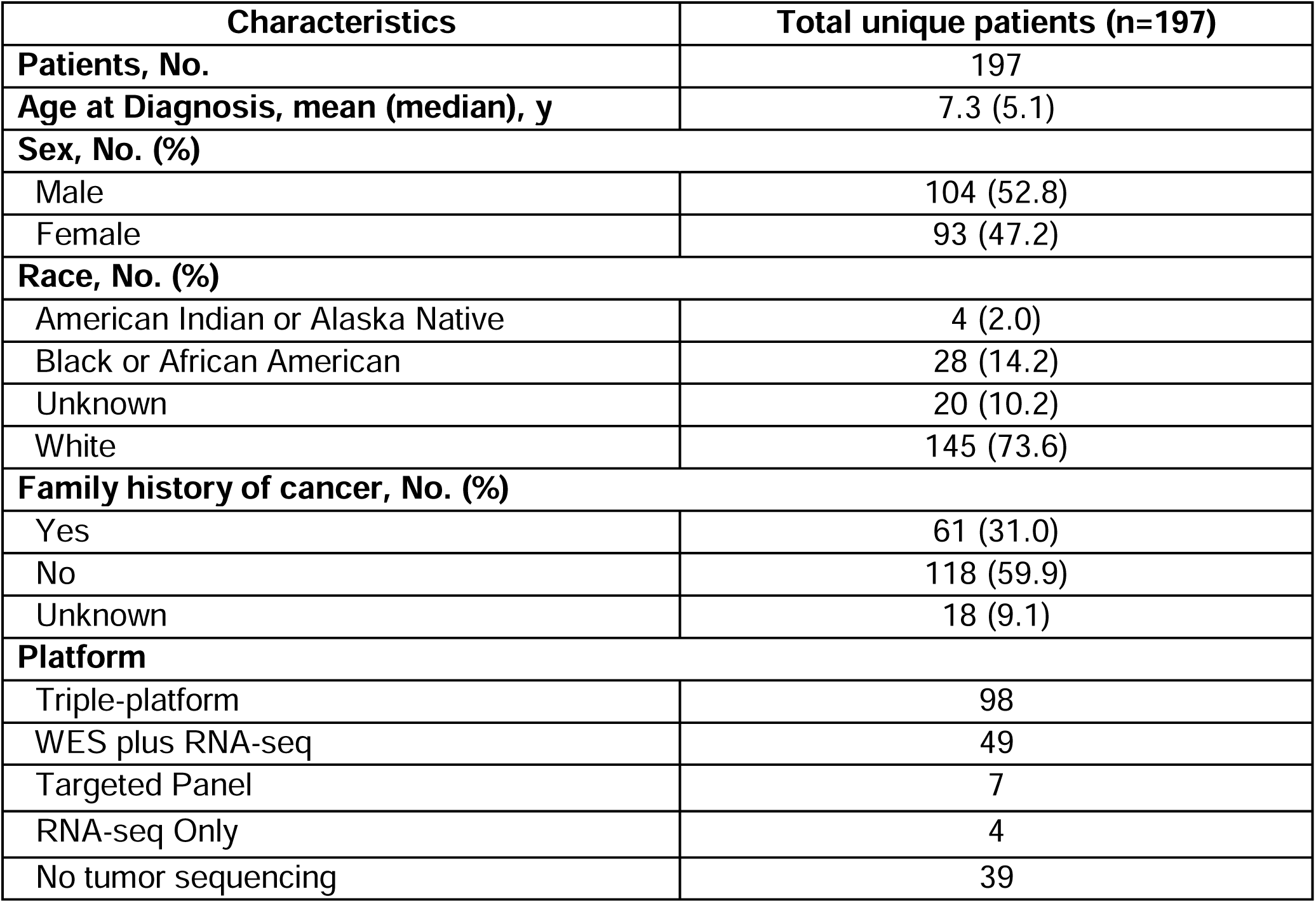
Clinical characteristics of patients with P/LP variants.

### The Distribution and Role of P/LP Variants in aoCPG and poCPG

The distribution of variant roles differed significantly between aoCPG and poCPG (Fisher’s exact test, P<.001; Figure 2; Supplementary Table 2). Among 66 aoCPG variants, 7 (10.6%) were PF, 19 (28.8%) ACMG-SF, 35 (53.0%) non-ACMG-SF, and 5 (7.6%) indeterminate. In contrast, among 87 poCPG variants, 77 (88.5%) were PF, 4 (4.6%) ACMG-SF, and 6 (6.9%) non-ACMG-SF. In a secondary patient-level analysis restricted to patients with autosomal dominant aoCPG or poCPG variants who underwent triple-platform or exome plus RNA sequencing, second somatic events were more frequently observed in poCPG than aoCPG (42 of 56 [75.0%] vs 7 of 51 [13.7%]; Pearson’s Chi-squared test with Yates continuity correction, P<.001).

**Figure 2.**
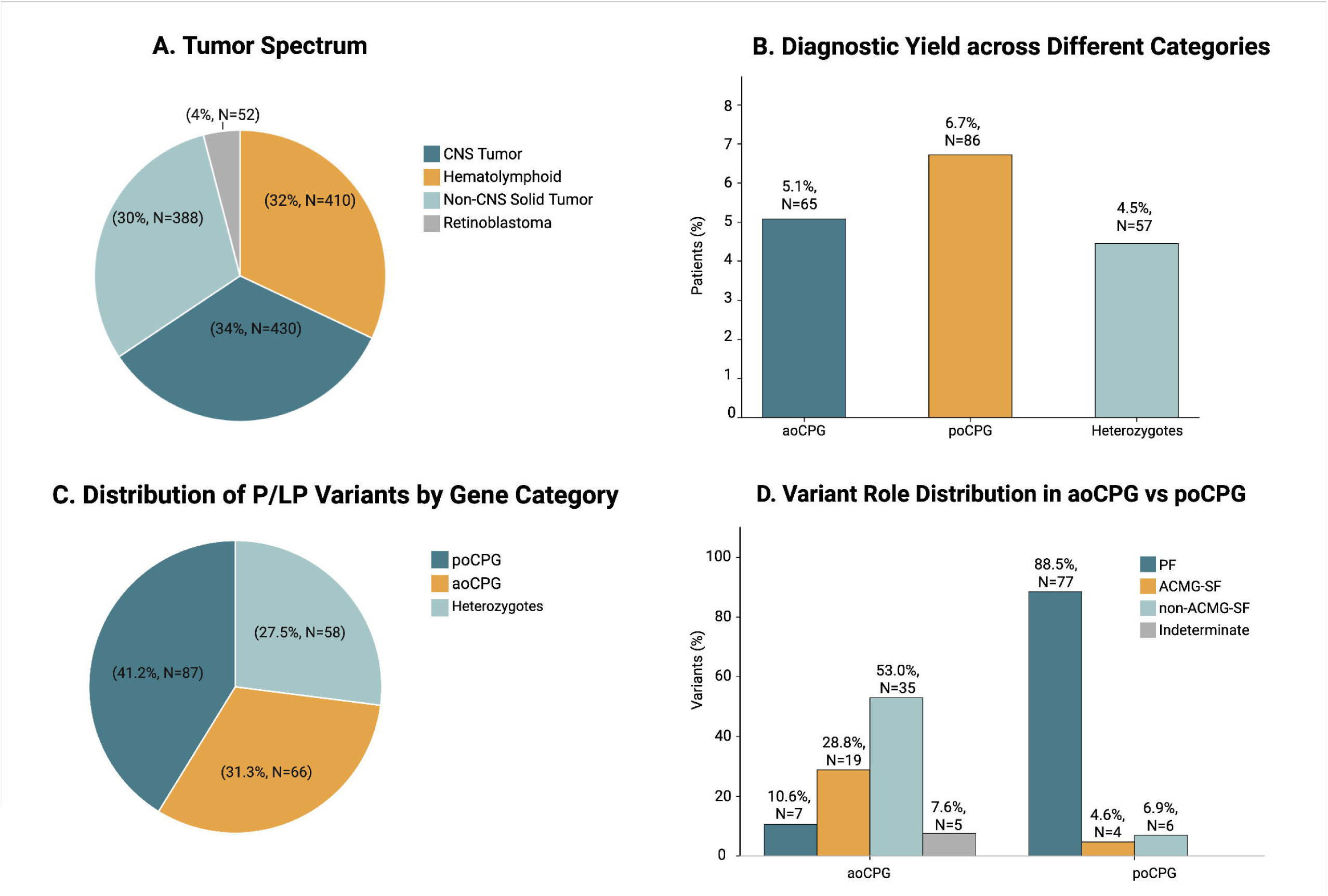
(A) Distribution of tumor types among 1,280 patients, including CNS tumors, hematolymphoid malignancies, non-CNS solid tumors, and retinoblastoma. (B) Diagnostic yield of germline P/LP variants across gene categories, showing the proportion of patients with at least 1 variant in aoCPG, poCPG, or as heterozygotes with variants in autosomal recessive genes. Category-specific yields are not mutually exclusive. (C) Distribution of P/LP variants (n=211) by gene category, including poCPG, aoCPG, and heterozygous variants in autosomal recessive genes. (D) Distribution of variant roles within aoCPG and poCPG categories, including primary findings (PF), secondary findings (ACMG-SF and non–ACMG-SF), and indeterminate variants.

### Clinical and molecular features of P/LP variants in aoCPG

The 66 aoCPG variants occurred in 65 patients, 53 of whom had tumor sequencing data available (Figure 3; Supplementary Figure 1; Supplementary Figure 2). Median age at diagnosis was 5.8 years. Seven variants (10.6%) were classified as PF based on concordant tumor phenotype and/or somatic findings. Three were classified as PF based on the tumor type and supporting somatic alterations: *CDKN2A* (n=1), *MLH1* (n=1), and *MSH6* (n=1). Both Lynch syndrome (LS) patients presented with high-grade glioma (HGG) and showed a second somatic hit, with a tumor mutational burden of 120 mutations/Mb (ultrahypermutated) and 4 mutations/Mb (low/borderline), respectively. Both tumors exhibited loss of MMR protein expression in tumor cells with retained stromal staining, supporting PF designation. One patient with PF in LS received immunotherapy but died of disease progression.

**Figure 3.**
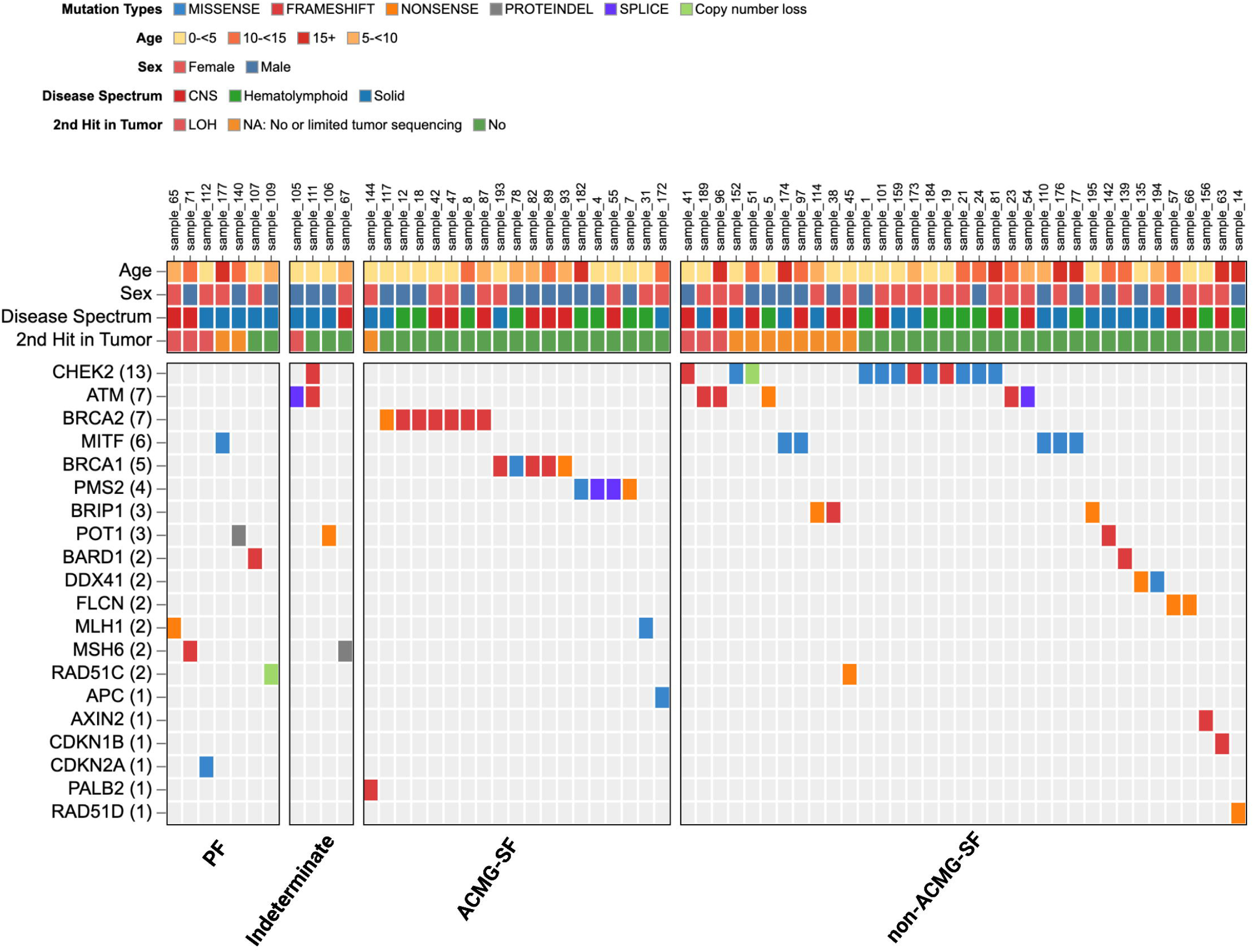
Integrated heatmap/oncoprint of P/LP variants in adult-onset cancer predisposition genes (aoCPGs) identified across the study cohort. Heatmap summarizing P/LP variants in aoCPGs identified across individuals, stratified by the distribution of variant roles (primary findings, secondary findings (ACMG-SF or non-ACMG-SF), or indeterminate findings). Columns represent individual participants and rows represent genes, ordered by frequency of alteration. Variant types are color-coded (missense, frameshift, nonsense, protein extension, splice site, and copy-number loss). Annotation tracks above the oncoprint indicate age at diagnosis, patient sex, disease spectrum, and availability of second-hit evidence. The number of patients harboring a variant in each gene is shown in parentheses.

*BARD1* and *RAD51C* variants were detected in patients with neuroblastoma and regarded as PF.^34–36^ Targeted tumor sequencing showed a single *BARD1* hit and retained protein expression in conjunction with a somatic (*ALK*) NM_004304.5:c.3522C>A p.(Phe1174Leu) variant; the patient received talazoparib, irinotecan, and lorlatinib but died of disease progression. No second somatic hit was identified in *RAD51C*, and no targeted therapy was administered. The tumor harbored a hemizygous loss of *ATRX*. Two additional variants were classified as PF based solely on tumor histology, as tumor sequencing data were unavailable. These included a *POT1* variant in a patient with osteosarcoma and Hodgkin lymphoma, and the founder variant (*MITF*) NM_198159.2:c.1255G>A p.(Glu419Lys) in a patient with melanoma.^37–39^

Five variants in four patients were classified as indeterminate, including one patient with co-occurring *ATM* and *CHEK2* variants. One LS patient with HGG harbored (*MSH6*) NM_000179.2:c.1618_1620del p.(Leu540del) with no tumor LOH; immunohistochemistry showed retained protein expression (that is expected for this variant type) and the tumor exhibited low TMB, supporting indeterminate classification. The patient received immunotherapy (pembrolizumab) but died due to disease progression. One patient with neuroblastoma harbored somatic *MYCN* amplification and a germline *ATM* variant associated with copy-neutral LOH at 11q, a region frequently altered in this tumor type. Another neuroblastoma patient harbored co-occurring germline *ATM* and *CHEK2* variants and somatic (*ALK*) NM_004304.5:c.3824G>A p.(Arg1275Gln); the contribution of either *ATM* or *CHEK2* to tumorigenesis remained uncertain.^35,40^ A *POT1* variant in a neuroblastoma patient was considered indeterminate, given emerging but limited evidence for *POT1* in neuroblastoma predisposition.^41^ No additional somatic variants were detected in the tumor sample.

The remaining 54 (81.8%) aoCPG variants were classified as SF, including 19 (28.8%) ACMG-SF and 35 (53.0%) non-ACMG-SF. Among the ACMG-SF, five LS patients (4 *PMS2* and 1 *MLH1*) with available tumor data showed no somatic features of MMR deficiency, such as a second hit or elevated TMB. Instead, these tumors harbored well-established somatic drivers. The tumor profiles of the four patients with *PMS2* variants included pleomorphic xanthoastrocytoma with an ALK kinase fusion and homozygous *CDKN2A* deletion; acute myeloid leukemia (AML) with a *KAT6B::NUTM2B* fusion transcript; AML with monosomy 7 and somatic variants in *ASXL1*, *EZH2*, *NRAS*, and *PPM1D* detected by targeted sequencing; and T-lymphoblastic leukemia/lymphoma (TLL) with expected somatic drivers in *NOTCH1* and *CDKN2A*.^42^ The remaining LS patient with an *MLH1* variant presented with B-lymphoblastic leukemia/lymphoma (BLL) driven by an *ETV6::RUNX1* fusion transcript. Of note, in patients with heterozygous *PMS2* variants, the possibility of additional alterations in exons 11–15 could not be fully excluded because of the high sequence homology in this region. However, the absence of somatic features suggestive of MMR deficiency, including elevated TMB, together with the presence of expected tumor drivers, argues against this possibility.

Thirteen P/LP variants in homologous recombination repair (HRR) genes, including *BRCA1* (n = 5), *BRCA2* (n = 7), and *PALB2* (n = 1), were classified as ACMG-SF, with tumor sequencing data available for 11 patients. These P/LP variants in HRR genes were observed across multiple tumor types with somatic tumor drivers. For example, *BRCA1/BRCA2* variants were detected in patients with low-grade gliomas harboring tyrosine kinase fusions (e.g., *KIAA1549::BRAF*), as well as embryonal tumors, including atypical teratoid/rhabdoid tumors with somatic *SMARCB1* alterations and medulloblastoma (Supplementary Table 1). *BRCA1/BRCA2* variants were also identified in a subset of patients with hematolymphoid malignancies, including BLL with *KMT2A* rearrangement or *ETV6::RUNX1* fusion transcript, AML with a *DEK::NUP214* fusion transcript, and Burkitt’s lymphoma with an *IGH::MYC* rearrangement. The (*APC*) NM_000038.6:c.3920T>A p.(Ile1307Lys) variant occurred in one patient with papillary thyroid carcinoma co-occurring with a *PTEN* variant, a known driver of this tumor type.

Thirty-five variants across 12 genes were classified as non-ACMG-SF, with tumor sequencing data available for 27 patients. Of these, 23 had expected somatic drivers (e.g., *CRLF2* position-effect; Supplementary Table 1). This group included 23 variants in 6 HRR genes (*ATM*, *BARD1*, *BRIP1*, *CHEK2*, *RAD51C*, and *RAD51D*), which confer low-to-moderate penetrance cancer risk. *CHEK2* accounted for 13 variants, including 2 founder alleles in 5 cases, NM_007194.4:c.470T>C p.(Ile157Thr) and c.1100del p.(Thr367MetfsTer15), both with gnomAD allele frequencies >0.1%. The c.1100del variant occurred in a high-grade neuroepithelial tumor with an *MN1::PATZ1* fusion resulting from a focal 22q deletion that also involved *CHEK2*, supporting non-ACMG-SF classification despite increased tumor VAF. The remaining *CHEK2* variants were observed across diverse malignancies, including BLL with distinct genetic subtypes (e.g., hyperdiploid and Philadelphia-like), AML with *NPM1* alterations, TLL, low-grade glioma, medulloblastoma, Ewing sarcoma with *EWSR1::ERG* fusion, and melanoma with *BRAF* p.(Val600Glu).

*ATM* variants were detected in medulloblastoma non-WNT/non-SHH, Wilms tumor with somatic *DROSHA* alterations, glioma with *H3F3A* alterations, and BLL harboring somatic *KRAS*, *LEF1*, and *CDKN2A* alterations. An additional *BARD1* variant was identified in dermatofibrosarcoma protuberans with a *COL1A1::PDGFB* fusion transcript and retained *BARD1* protein expression by immunohistochemistry in the tumor sample. A *RAD51D* variant was identified in BLL with a *NUP214::ABL1* fusion transcript. Heterozygous *BRIP1* was detected in a patient with Wilms tumor harboring a germline *WT1* variant with biallelic inactivation. Additional founder variants included (*MITF*) NM_198159.2:c.1255G>A p.(Glu419Lys) (n=5), detected in patients without melanocytic tumors. The remaining variants involved *AXIN2*, *CDKN1B*, *DDX41* (n=2), *FLCN* (n=2), and *POT1*. *AXIN2* was identified in a patient with multiple tumor types, including AML, melanoma, meningioma, and rhabdoid tumor. *CDKN1B* was identified in myxopapillary ependymoma; *DDX41* in rhabdomyosarcoma with somatic HRAS alterations and in a patient with a history of Wilms tumor and germinoma; and *FLCN* in HGG with somatic *BRAF* or *H3F3A* alterations.

### Clinical and Molecular Features of P/LP Variants in poCPG

Unlike aoCPG, poCPG variants were usually supported by a syndrome-concordant tumor phenotype and often by tumor-based molecular corroboration. As expected, P/LP variants in poCPG were predominantly classified as PF and none were indeterminate. Of 211 P/LP variants, 87 (41.2%) occurred in 26 poCPGs and were detected in 86 patients, 58 of whom had tumor sequencing data available. The median age at diagnosis was 4.1 years. Overall, 77 of 87 (88.5%) were PF because the tumor type or clinical presentation was concordant with the expected gene-associated phenotype (Figure 4; Supplementary Figure 3; Supplementary Figure 4). PF classification was further supported by tumor evidence of a second hit in many cases, including LOH (n=28), a second somatic variant (n=8), hypermutation in biallelic *PMS2*-associated constitutional mismatch repair deficiency (CMMRD, n=1), and targeted DNA evidence of LOH or a second somatic event in *NF2* and *CBL* (n=2). Germline CNVs involving *RB1* (n=3), *NF2* (n=1), *PAX5* (n=1), and *WT1* (n=1) were identified in 6 patients, 5 with corresponding somatic second-hit events.

**Figure 4.**
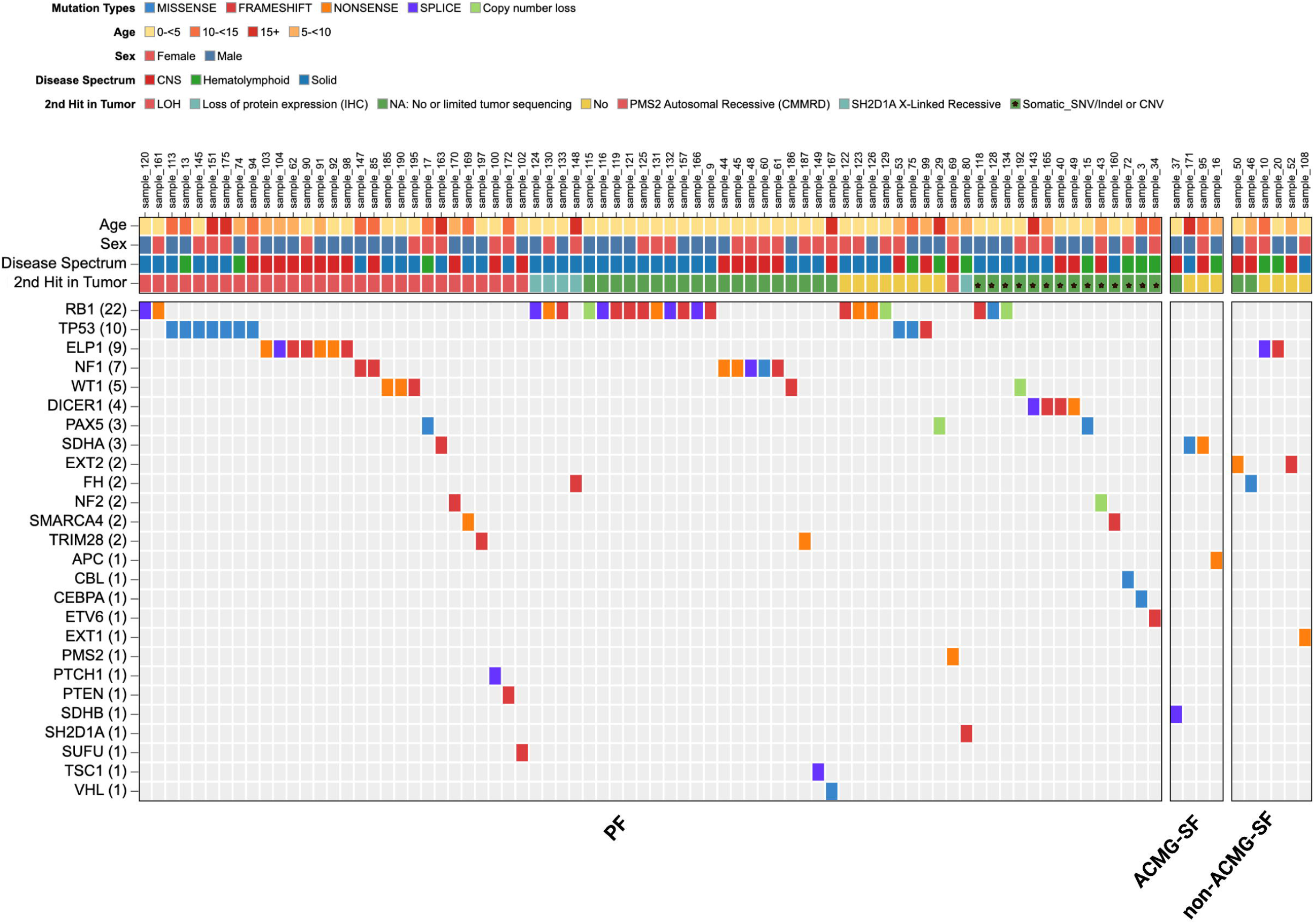
Integrated oncoprint of pathogenic or likely pathogenic variants in pediatric-onset cancer predisposition genes (poCPGs) identified across the study cohort. Oncoprint summarizing pathogenic and likely pathogenic variants in poCPGs identified across individuals, stratified by the distribution of variant roles (primary findings, secondary findings (ACMG-designated or non-ACMG), or indeterminate findings), and highlighting the distribution and spectrum of clinically actionable variants detected across cancer predisposition and medically actionable genes. Columns represent individual participants and rows represent genes, ordered by frequency of alteration. Variant types are color-coded (missense, frameshift, nonsense, protein extension, splice site, and copy number loss). Annotation tracks above the oncoprint indicate age at diagnosis, patient sex, disease spectrum, and availability of second-hit evidence. The number of patients harboring a variant in each gene is shown in parentheses.

Additional poCPG variants were classified as PF based on syndrome-concordant tumor type, clinicopathologic features, and/or immunohistochemistry when a second somatic event was not demonstrated or tumor sequencing was unavailable. Examples included *FH-*deficient renal cell carcinoma, *TP53*-associated tumors, *RB1*-associated retinoblastoma, *PAX5*-associated B-lymphoblastic leukemia/lymphoma, *WT1*- or *TRIM28*-associated Wilms tumor, *NF1*-associated optic pathway/hypothalamic glioma, and *SH2D1A*-associated Burkitt lymphoma. Two patients with *NF1* received MEK inhibitor and one with *NF2* received bevacizumab.

Ten variants (11.5%) were classified as SF, including 4 (4.6%) ACMG-SF and 6 (6.9%) non-ACMG-SF. The ACMG-SF in poCPG included *SDHA* variants in Ewing sarcoma with an *EWSR1* rearrangement and in *SHH*-activated medulloblastoma with somatic *TP53* alteration, *SDHB* in ependymoma with a *ZFTA::RELA* fusion transcript, and *APC* in hypodiploid BLL. The non-ACMG-SF in poCPG comprised *ELP1* variants in AML with *NPM1* alterations and in BLL with a *BCR::ABL1* fusion transcript; *EXT2* in craniopharyngioma with a somatic *CTNNB1* alteration; *EXT1* in neuroblastoma with a somatic *TERT* alteration and in dysembryoplastic neuroepithelial tumor (DNET) with an *FGFR1* tandem duplication; and *FH* in HGG with a *FOXR2* position effect.^43^

### Clinical and Molecular Features of P/LP Variants in Heterozygotes

Of 211 P/LP variants, 58 (27.5%) were heterozygous in autosomal recessive genes. *MUTYH* accounted for 23, including 21 founder alleles: NM_001128425.1:c.1187G>A p.(Gly396Asp), and NM_001128425.1:c.536A>G p.(Tyr179Cys). One case with (*MUTYH*) NM_001128425.1:c.1187G>A p.(Gly396Asp) and an oncocytic renal neoplasm was classified as indeterminate due to tumor LOH without additional pathogenic alterations; heterozygous *MUTYH* variants with somatic second hits have been reported in adult and pediatric tumors but require further study.^4,44^ Two additional cases demonstrated LOH: one involving *NBN* in a poorly differentiated carcinoma harboring somatic *TP53* alterations and aneuploidy, and another involving *NTHL1* in adrenocortical carcinoma, a tumor type typically characterized by genomic stability.^45^

## Discussion

In this consecutive cohort of 1,280 pediatric patients with cancer undergoing broad germline sequencing, 197 (15.4%) harbored 211 P/LP variants, consistent with prior reports (8%–18%).^8–11^ Stratification by gene category showed that 65 patients (5.1%) had variants in aoCPG, 86 (6.7%) in poCPG, and 57 (4.5%) were heterozygotes of autosomal recessive genes. Although nearly one-third of P/LP variants (66 of 211) occurred in aoCPG, only 7 (10.6%) were classified as PF, whereas 54 (81.8%) were SF. In contrast, the 77 (88.5%) variants in poCPG were PF. These findings underscore that, in pediatric oncology, pathogenicity in aoCPG does not equate to tumor relevance.

Interpreting the role of germline variants beyond the pathogenic classification is strengthened by integrating clinical context with tumor phenotype, immunohistochemistry, and somatic genomic data in the pediatric oncology setting.

These findings align with prior reports suggesting that aoCPG confer lower cancer risk in children and adolescents and show that most such variants are not clearly implicated in the presenting tumor when interpreted with clinicopathologic and molecular evidence.^2^ This distinction has direct clinical implications. Pretest counseling should prepare families for the possibility that broad germline sequencing may identify medically relevant adult-onset predisposition variants that are unrelated to the child’s current cancer but still carry implications for future surveillance, family counseling, and cascade testing.

In our cohort, PF and indeterminate findings indicate that a subset of adult-onset variants may still be clinically relevant in selected cases, particularly when supported by a concordant tumor phenotype and corroborating molecular evidence. Similar to the poCPG, a few adult-onset variants were informative for considering targeted therapy.

This was illustrated by LS, which was identified in 8 of 1,280 patients (0.6%); however, only 2 tumors showed somatic and immunophenotypic evidence of mismatch repair deficiency consistent with PF, 1 case was classified as indeterminate, and only 2 patients received immunotherapy.^3^ Thus, LS in a child with cancer does not necessarily indicate a causal role and should not, in isolation, guide immunotherapy decisions without supporting histopathology and somatic evidence.^46^ Similarly, monoallelic variants in HRR genes are unlikely to confer sensitivity to poly (ADP-ribose) polymerase inhibitor (PARPi) therapy.^47,48^ In our cohort, *BARD1* variants were identified in neuroblastoma and dermatofibrosarcoma protuberans, and the neuroblastoma case was treated with a PARP inhibitor; however, emerging data suggest that biallelic *BARD1* loss may be required for therapeutic vulnerability.^6^ More broadly, these findings highlight the need to define when second-hit events are necessary for targeted treatment strategies and caution against overinterpreting isolated LOH, as illustrated by *CHEK2* in a tumor with an *MN1::PATZ1* fusion.

Our data also highlight the need to refine how PF and SF terminology are applied in pediatric oncology. Current ACMG-SFWG guidance is primarily intended for germline WGS/WES reporting and does not fully address the interpretive challenges that arise when broad cancer-predisposition panels are used in the evaluation of children with cancer. A reporting framework that incorporates test indication, tumor diagnosis/histopathology, immunohistochemistry, and somatic evidence would better distinguish variants that are PF from those that are truly SF. Such standardization could improve reporting clarity, post-testing counseling, cascade testing, and, in selected cases, access to targeted therapy or clinical trials.

This study has several limitations. First, tumor sequencing was unavailable for 31 patients with P/LP variants in aoCPG and poCPG, which may have reduced the ability to identify supportive somatic evidence and could have contributed to secondary or indeterminate classification in some cases. Second, because no age-matched pediatric control cohort was included, these data describe prevalence and clinical relevance within a sequenced clinical cohort rather than enrichment relative to the general pediatric population. Third, the assay was not clinically validated to detect all forms of second hits, including promoter methylation/epigenetic silencing events, and genomic scar and mutation signatures were not assessed.^4,49^ Fourth, classification of adult-onset vs pediatric-onset predisposition genes and classification of primary, secondary, and indeterminate findings necessarily relied on current evidence and may evolve as disease-gene associations are refined. Finally, these findings should be interpreted in the context of a single tertiary pediatric cancer referral center with broad clinical sequencing access and may not generalize fully to community-based settings or cohorts with less comprehensive tumor profiling.

In conclusion, broad germline testing in pediatric oncology frequently reveals P/LP variants in both aoCPG and poCPG, underscoring the importance of defining their roles beyond pathogenic classification. Integrating tumor diagnosis with histopathologic and molecular profiling, along with refining PF/SF terminology, is therefore critical for appropriate genetic counseling and for identifying the findings that may have therapeutic relevance.

## Data and code availability

All data supporting the findings of this study are available within the article and its supplemental information.

## Data Availability

All data supporting the findings of this study are available within the article and its supplemental information. Only deidentified data are provided.

## Acknowledgments

The authors thank the patients and their families at SJCRH for their participation. We also thank the genetic counselors and providers in the Cancer Predisposition Clinic, as well as members of the Molecular Pathology Laboratory and Clinical Genomics team at SJCRH. Preliminary findings of this study were presented at the 2026 ACMG Annual Clinical Genetics Meeting.

## Author Contributions

Conceptualization: M.K.E., J.L.M, K.E.N, J.M.K.; formal analysis: J.L.M., M.K.E.; data curation: J.L.M., M.K.E. V.P., T.C.; statistical analysis: C.W., C.L.; visualization: J.L.M., M.K.E.; writing – original draft: J.L.M., M.K.E.; writing – review & editing: J.L.M, V.P., T.C., M.C., P.R.B., M.R.W., S.K., C.W., C.L., L.W., K.E.N., J.M.K., M.K.E.

## Ethics Declaration

The St. Jude Children’s Research Hospital Institutional Review Board reviewed this study and determined it to be exempt under protocol 24-1762. Informed consent was not required by the IRB because the study used de-identified clinical and molecular data. All study procedures involving human participants were conducted in accordance with institutional requirements and applicable ethical standards.

## Web resources

HGVS, https://hgvs-nomenclature.org/

VariantValidator, https://www.variantvalidator.org/

Biorender, https://www.biorender.com/

**Additional Information**

**Supplementary Table 1.**

**Supplementary Table 2.**

**Supplementary Figures**

