## Supplementary Figures for "Prevalence and Clinical Significance of Adult-Onset Cancer Predisposition Variants in Pediatric Oncology"

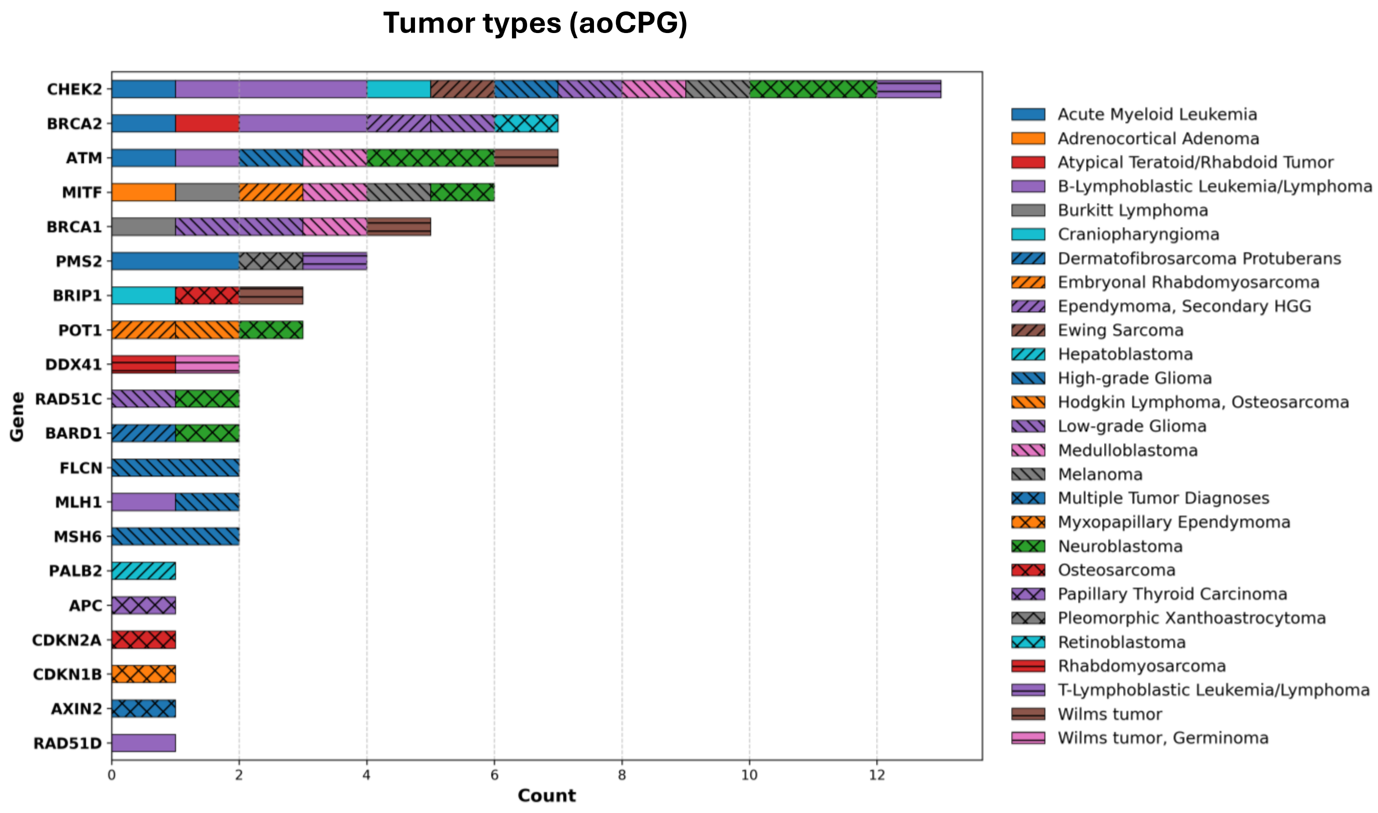


**Supplementary Figure 1. Tumor types/diagnoses of pathogenic or likely pathogenic variants in adult-onset cancer predisposition genes (aoCPGs).** Distribution of tumor diagnoses among patients with pathogenic or likely pathogenic variants in aoCPGs. Tumor types include hematologic, central nervous system, and solid malignancies, with some patients presenting with multiple malignancies.

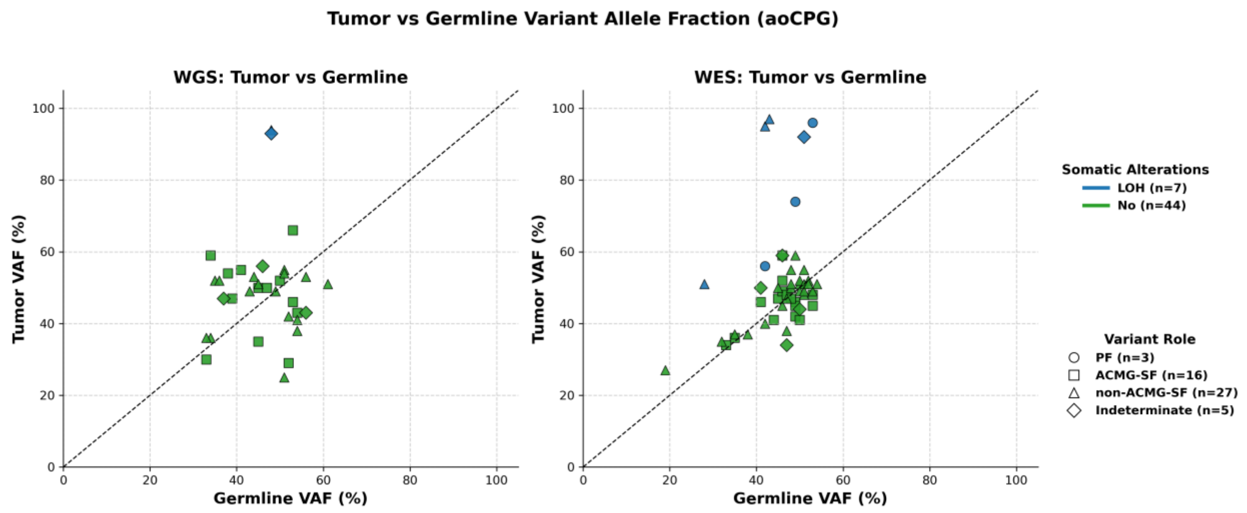


**Supplementary Figure 2. Tumor-germline variant allele fractions of pathogenic or likely pathogenic variants in adult-onset cancer predisposition genes (aoCPGs).** Comparison of tumor and germline variant allele fractions (VAFs) for autosomal dominant aoCPG variants identified by whole-genome sequencing (left) and whole-exome sequencing (right). Only cases with autosomal dominant germline SNV/indels are shown for which tumor whole-genome sequencing and/or whole exome sequencing are available. Each point represents an individual variant; point shapes indicate variant role classifications, and colors indicate presence or absence of somatic alterations, including loss of heterozygosity. The diagonal reference line denotes expected correspondence between tumor and germline VAFs. The majority of variants (n=43) did not show a significant difference in VAF between the tumor and germline; 7 tumor samples demonstrated LOH as defined by an increased VAF of at least 20%.

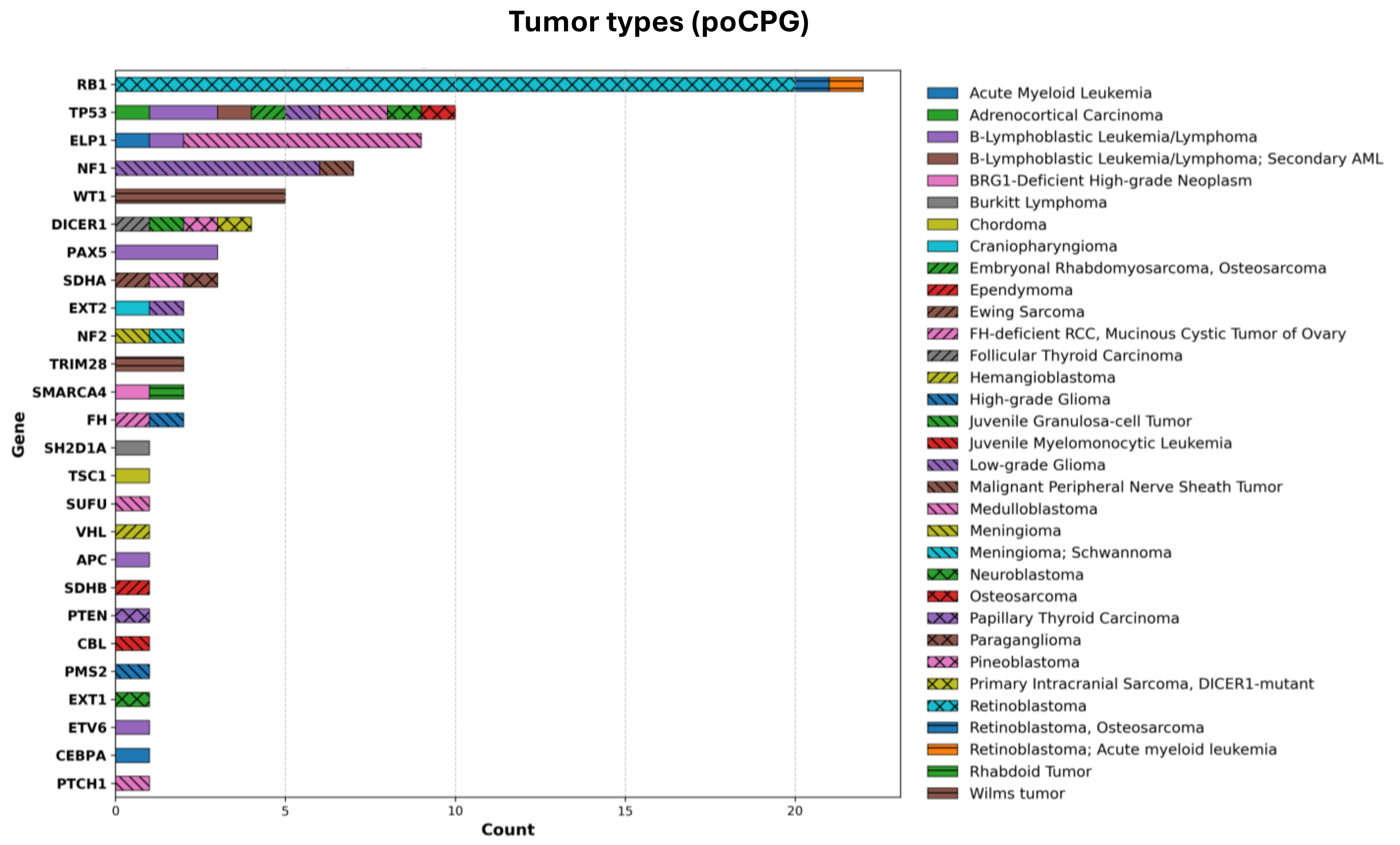


**Supplementary Figure 3. Tumor types and diagnoses of pathogenic or likely pathogenic variants in pediatric-onset cancer predisposition genes (poCPGs).** Distribution of tumor diagnoses among patients with pathogenic or likely pathogenic variants in poCPGs. Tumor types include hematologic, central nervous system, and solid malignancies, with some patients presenting with multiple malignancies.

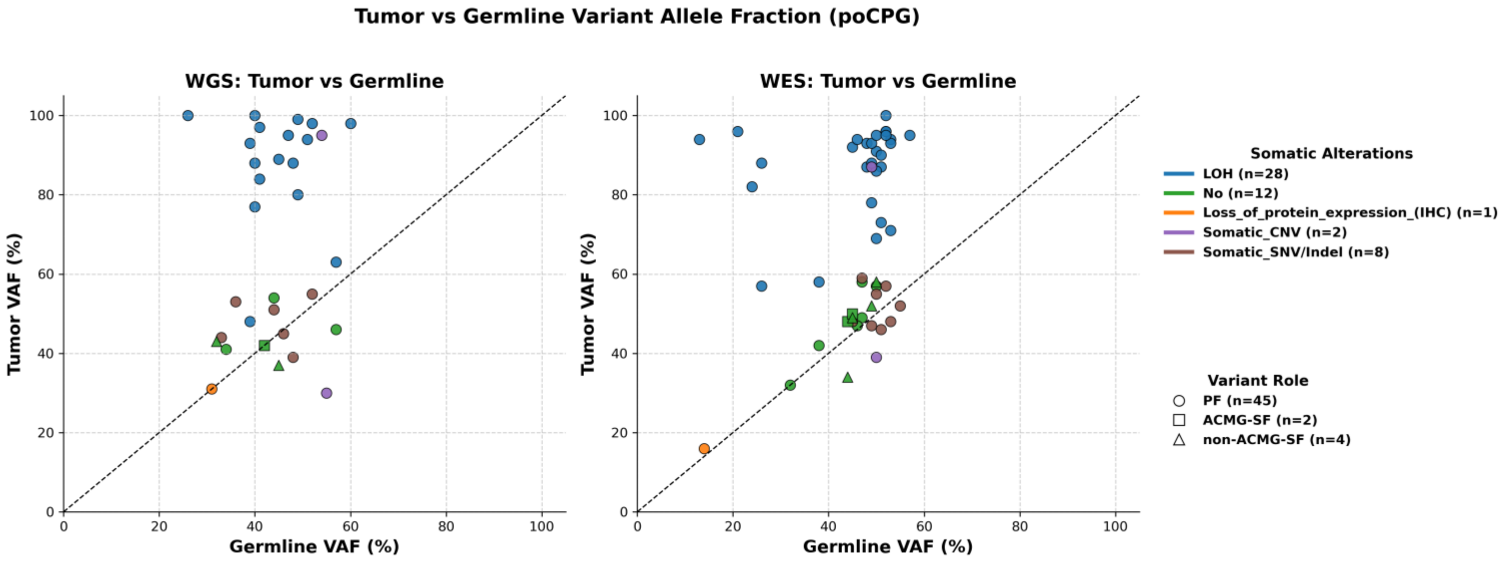


**Supplementary Figure 4**. **Tumor-germline variant allele fractions of pathogenic or likely pathogenic variants in pediatric-onset cancer predisposition genes (poCPGs).** Comparison of tumor and germline variant allele fractions (VAFs) for poCPG variants identified by whole-genome sequencing (left) and whole-exome sequencing (right). Only cases with autosomal dominant germline SNV/indels are shown for which tumor whole-genome sequencing and/or whole exome sequencing are available. Each point represents an individual variant; point shapes indicate variant role classifications, and colors indicate presence or absence of somatic alterations, including loss of heterozygosity. The diagonal reference line denotes expected correspondence between tumor and germline VAFs. In contrast to aoCPGs (Figure 3), the majority of variants (n=28) did demonstrate LOH as defined by an increased tumor VAF of at least 20%. In addition, second somatic hits were observed in 10 tumors, and loss of protein expression by IHC was observed in one tumor supporting second hit despite no second somatic variant was detected. Please note cases wither germline CNVs, autosomal recessive *PMS2* and *SH2D1A* X-Linked Recessive are not displayed.
